# Diagnostic Accuracy of GPT Multimodal Analysis on USMLE Questions Including Text and Visuals

**DOI:** 10.1101/2023.10.29.23297733

**Authors:** Vera Sorin, Benjamin S. Glicksberg, Yiftach Barash, Eli Konen, Girish Nadkarni, Eyal Klang

## Abstract

**Objective:** Large Language Models (LLMs) have demonstrated proficiency in free-text analysis in healthcare. With recent advancements, GPT-4 now has the capability to analyze both text and accompanying images. The aim of this study was to evaluate the performance of the multimodal GPT-4 in analyzing medical images using USMLE questions that incorporate visuals.

**Methods:** We analyzed GPT-4’s performance on 55 USMLE sample questions across the three steps. In separate chat instances we provided the model with each question both with and without the images. We calculated accuracy with and without the images provided.

**Results:** GPT-4 achieved an accuracy of 80.0% with images and 65.0% without. No cases existed where the model answered correctly without images and incorrectly with them. Performance varied across USMLE steps and was significantly better for questions with figures compared to graphs.

**Conclusion:** GPT-4 demonstrated an ability to analyze medical images from USMLE questions, including graphs and figures. A multimodal LLM in healthcare could potentially accelerate both patient care and research, by integrating visual data and text in analysis processes.

## Introduction

Large Language Models (LLMs) such as ChatGPT by OpenAI have shown impressive skills in free-text analysis and generation in various tasks related to healthcare^1-6^. Numerous publications detail LLMs’ performance on the United States Medical Licensing Examination (USMLE), with GPT-4 achieving 80-90% success rate^7^. Performance however was evaluated on textual data only, either excluding questions with visuals, or including them without the associated images.

While these abilities of LLMs are remarkable, language analysis is only one aspect of patient care. Diagnosis and management rely not only on anamnesis and clinical history, but include physical examination, and supplemental tests such laboratory, ECG, imaging and pathology images^8,9^. These types of data can include images, audio, and video.

GPT-4 by Open-AI, can now analyze images alongside textual data^10^. This advancement brings the algorithm one step closer to artificial general intelligence (AGI). In healthcare, it may enable more comprehensive and practical medical care, with many additional potential uses. The aim of our study was to evaluate GPT-4 performance in analyzing medical images, specifically using USMLE questions that incorporate visuals.

## Methods

### Experimental Design

We evaluated OpenAI’s GPT-4 multimodal model on a set of 55 USMLE sample questions. These questions were sourced from sample tests for steps 1, 2CK, and 3, available at the USMLE official website. The same sample was used in a publication by Nori et al. (Microsoft and OpenAI) evaluating GPT-4 performance on the USMLE^11^. As they have been extensively studied before, we did not analyze any questions that were text-only. Instead, we used all questions that included images and conducted two sets of experiments: one where the model was provided with both text and accompanying images, and another including only the text without images.

Questions were further divided into two groups based on the type of accompanying images - those with graphs versus those with figures. All interactions with the model were carried out using OpenAI’s web interface.

### Prompts

Two distinct prompt types were used to elicit responses from the model:

> *“Please answer the following USMLE question based on the attached image”*.
>
> *“The following USMLE question is based on an image that is not provided. Please answer the question to the best of your ability using the information available*.*”*

While both prompts incorporated the text from the questions, only the first prompt included the images alongside the text. Each prompt type was assessed in a separate chat instance for every question.

### Statistical Analysis

Accuracy was the primary metric for evaluating the model’s performance, calculated as the percentage of correctly answered questions over the total number of questions in each category, and separately for the same set of questions answered with images and without the images provided. A Fisher’s exact test was employed to assess the statistical significance of the difference in performance between questions answered with and without images, and for questions with figures versus questions with graphs.

All data preprocessing and statistical analyses were conducted using Python version 3.10, with Pandas for data manipulation and SciPy for statistical testing.

## Results

### Dataset Description

The dataset comprised 55 USMLE sample questions distributed over three examination steps: 29 from Step 1, 13 from Step 2CK, and 13 from Step 3 (**Figure 1**). Questions accompanied by graphs were fewer: 11/29 in Step 1, 1/13 in Step 2CK, and 3/13 in Step 3.

**Figure 1:**
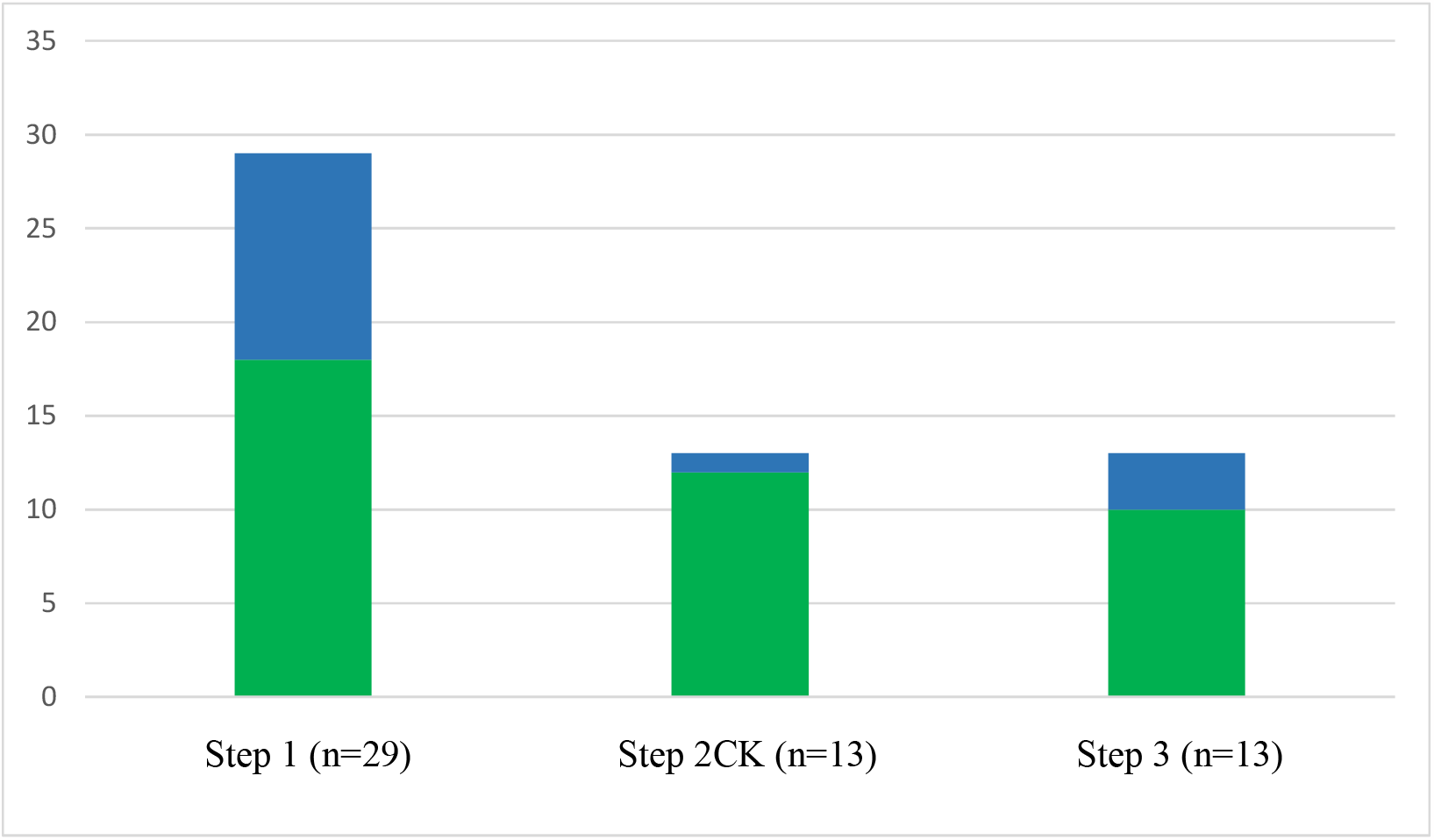
Distribution of sample USMLE Questions with images (n=55). Questions with graphs are represented in blue, questions with figures are in green.

### Overall Performance

GPT-4 achieved an overall accuracy of 80.0% (44/55 correct answers) when images were provided and 65.0% (36/55 correct answers) without them (p=0.089). There was no scenario when the model answered a question correctly when an image was not provided and then incorrectly with the image. When looking at the correctly answered questions when images were not provided, it is interesting to understand the “reasoning” of the model, where in some cases the model “chose” the correct answer based on overall probability, or where there were enough clinical details/image description that enabled the correct diagnosis. Example cases are provided in the **Supplement**.

### Performance by USMLE Step

GPT-4’s accuracy differed across the three steps, both with and without the accompanying images (**Table 1**). The model achieved 100% accuracy (13/13 correct answers) on Step 3 questions when images were provided. The model’s performance dipped to 55.2% (16/29 correct answers) for Step 1 questions without the images provided.

**Table 1:** Accuracy by USMLE Step.

| Exam Step | Accuracy with Image (%) | Accuracy without Image (%) | p Value |
| --- | --- | --- | --- |
| Step 1 | 20/29 (69.0) | 16/29 (55.2) |  |
| Step 2CK | 11/13 (84.6) | 10/13 (76.9) |  |
| Step 3 | 13/13 (100.0) | 10/13 (76.9) |  |
| Overall | 44/55 (80.0) | 36/55 (65.5) | 0.089 |

### Performance by Image Type

When comparing performance in questions including graphs versus questions with figures, overall performance was better in questions with figures (with or without the images). When evaluating questions with graphs, accuracy was 20% (can be considered random choices) when images were not provided but improved to 40% when images were included (**Table 2**).

**Table 2:** Accuracy by Image Type.

| Image Type | Accuracy with Image (%) | Accuracy without Image (%) | p Value |
| --- | --- | --- | --- |
| Figures | 38/40 (95.0) | 33/40 (82.5) | <0.001 |
| Graphs | 6/15 (40.0) | 3/15 (20.0) | <0.001 |

## Discussion

In this study we evaluated the performance of multimodal GPT-4 using USMLE questions that incorporate visuals. There are several important findings: (1) Overall accuracy on USMLE questions reached 80% when images were provided. This is close to the previously reported accuracy of 80-90% based on text analysis alone^7^. (2) The model’s accuracy improved with the inclusion of images, though the difference was not statistically significant. (3) GPT-4 performed better on questions with figures compared to graphs, but its accuracy in questions with graphs improved when images were provided.

The 80% accuracy of GPT-4 on USMLE questions with visuals was slightly lower than the reported 83.76% on the complete set of questions as noted by Nori et al. (including questions with visuals based on textual data only)^11^. When examining each step individually, performance on Step 3 questions with visuals was 100%, surpassing Nori et al.’s reported 81.25% on the complete Step 3 set^11^. Accuracy overall improved when images were provided. Despite the lack of statistical significance for the difference in performance, no questions were answered correctly without images and then incorrectly with them. When answers were correct, the model correctly described and interpreted the images, underscoring its capability to analyze medical visuals.

The potential of a multimodal LLM in healthcare is vast. As patient care relies on diverse data sources, the ability to incorporate both visual and text data has broad applications. Such models can evaluate patient notes alongside radiology or pathology images and interpret findings from physical exams alongside patient medical history and anamnesis. It is interesting to note that GPT-4 performed well on some questions even without the images provided. This enhances the fact that clinical history is an essential part of medical diagnosis ^12,13^.

Multimodal LLMs could also impact research. The ability to analyze graphs and figures alongside text could enhance data collection abilities, and literature reviews. It can reduce human bias in data collection and interpretation, and potentially even provide insights from complex datasets.

This study has several limitations. The sample size was small, as we aimed for homogeneous questions that will be as close to the real examination as possible, utilizing only the official USMLE sample test and excluding other medical question banks. This set of questions was also used by Nori et al. from Microsoft and OpenAI^11^. Questions from Steps 2 and 3, as well as those with graphs, were underrepresented. Additionally, since the questions are publicly available online, there’s potential for training contamination^11^.

To conclude, GPT-4 demonstrated an ability to analyze medical images from USMLE questions, including graphs and figures. A multimodal LLM in healthcare could potentially accelerate both patient care and research, by integrating visual data and text in analysis processes.

## Data Availability

All data produced in the present study are available upon reasonable request to the authors

